# Exploring Zero-Shot Cross-Lingual Biomedical Concept Normalization via Large Language Models

**DOI:** 10.1101/2025.02.27.25323007

**Authors:** Hossein Rouhizadeh, Anthony Yazdani, Boya Zhang, Douglas Teodoro

## Abstract

Over the past few years, discriminative and generative large language models (LLMs) have emerged as the predominant approaches in natural language processing. However, despite significant advancements, there remains a gap in comparing the performance of discriminative and generative LLMs in cross-lingual biomedical concept normalization. In this paper, we perform a comparative study across several LLMs on the challenging task of cross-lingual biomedical concept normalization via dense retrieval. We utilize the XL-BEL dataset covering 10 languages to evaluate the model’s capacity to generalize across various linguistic contexts without further adaptation. The experimental findings demonstrate that e5, a discriminative model, exhibited superior performance, whereas BioMistral emerged as the top-performing generative LLM. The code for reproducing the experiments is available at: https://github.com/hrouhizadeh/zsh_cl_bcn.

## 1. Introduction

Large language models (LLMs) have significantly advanced the field of biomedical natural language processing in recent years, impacting numerous applications from text generation to sentiment analysis [1]. In biomedical information extraction, LLMs have progressed in several long-standing tasks such as named entity recognition [2], adverse drug events normalization [3] relation extraction [4, 5], and concept normalization [6].

For large-scale concept normalization tasks, retrieval-based approaches are essential [4]. Rather than relying on traditional explicit term classification, these methods prioritize searching for the most relevant concepts. This is especially important in the biomedical domain, where terminologies comprise thousands of entities. Dense retrieval supported by LLMs represents a significant advancement in information retrieval, offering an advanced approach to searching large document collections and finding the most relevant documents for a given query [7, 8]. This method encodes queries and documents into dense vector representations, enabling models to capture semantic meaning beyond simple string matching.

Despite the increasing role of LLMs in biomedical concept normalization, their effectiveness in zero-shot cross-lingual concept normalization remains understudied. In this study, we aim to address this gap by investigating the performance of discriminative and generative LLMs in zero-shot, large-scale, cross-lingual concept normalization. We used XL-BEL [9], a cross-lingual biomedical concept normalization benchmark, to assess various discriminative and generative LLMs in this setting via a dense retrieval approach.

## 2. Methods

### 2.1. Resources

We used two main dataset resources in our zero-shot evaluation pipeline:

- The Unified Medical Language System (UMLS) is a comprehensive biomedical knowledge resource that integrates over 4 million concepts from more than 200 vocabulary sources. It organizes these concepts using unique identifiers called concept unique identifiers (CUIs), each representing a specific meaning and encompassing synonymous terms from various vocabularies. This study employs the UMLS 2022AB version as the primary knowledge base for concept normalization.
- XL-BEL [9] is a cross-lingual dataset for biomedical entity linking that spans 10 typologically diverse languages including Chinese, English, Finnish, German, Japanese, Korean, Russian, Spanish, Thai, and Turkish. The dataset only includes a test set of 1,000 instances for each language. We follow [9] and restrict UMLS to WikiMed’s subset of 62,531 CUIs, corresponding to 399,931 UMLS English concepts, as our target ontology.

### 2.2. Methodology

Our proposed system is designed to identify the most relevant UMLS concept for a given target term in 10 target languages. As shown in Figure 1, we first compute the cosine similarity between the input term (‘pulmonary emboli’) and UMLS concepts. Next, we select and order the top k concepts based on their similarity scores and associate them with their respective CUIs. In what follows, we describe each step:

**Figure 1.**
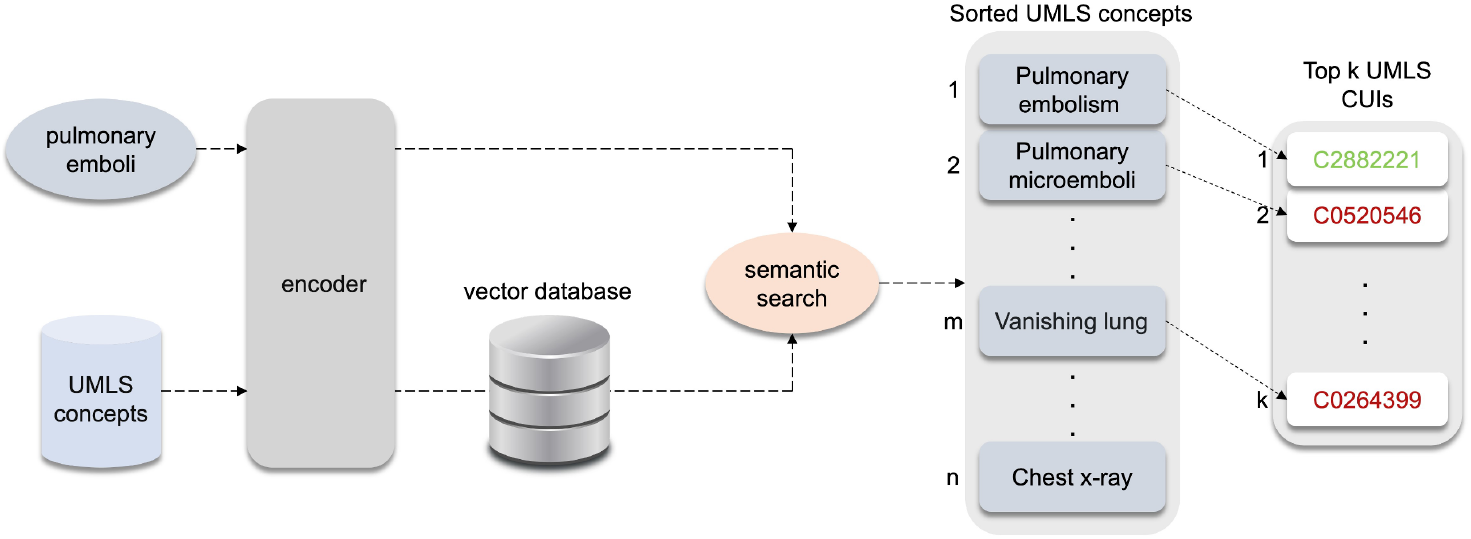
The overall performance of different models across 10 languages in the XL-BEL dataset.

Step 1: Computing UMLS concept embeddings: We begin by generating embeddings for UMLS concepts. This process involves inputting each concept into an LLM and extracting the [CLS] token from the final hidden layer as the concept’s representation. Given the extensive size of our target UMLS subset, efficient storage of the computed embeddings is crucial. To address this challenge, we utilize the Qdrant vector database, which enables efficient storage and retrieval of high-dimensional vectors.

Step 2: Processing input term and comparison with UMLS concepts: This step identifies and retrieves the UMLS concepts most closely related to a given input. For each input term, we extract its representative embedding by generating the [CLS] token from the LLM’s last hidden layer. We then compute the cosine similarity between the input term and the UMLS concept embeddings stored in the vector database.

Step 3: Generating the CUI ranking list: In the final step, we sort the top k most similar concepts, retrieved in the previous step, based on their cosine similarity values and associate them with their corresponding CUIs. This ranking list of CUIs reflects the most relevant matches for the input term. For CUIs associated with multiple concepts, we select the highest-scoring concept to represent that CUI.

## 3. Result

We considered four discriminative and four generative LLMs in our experiments based on whether they were trained using multilingual data and available as open source i) discriminative - BERT (bert-base-multilingual-cased) [10], DistilBERT (distilbertbase-multilingual-cased) [11], e5 (multilingual-e5-large) [12], and MPNet (paraphrasemultilingual-mpnet-base-v2) [13]; and ii) generative - Aya (aya-23-8B) [14], Mistral (BioMistral-7B) [15], Llama (Meta-Llama-3-8B)[16], and StableLM (stablelm-2-1 6b) [17]. We assess the models’ effectiveness using Recall@N, a metric that measures the presence of relevant documents, i.e., concepts in our case, within the top *N* retrieved results.

Figure 2 presents the comparative performance of various models. Our analysis reveals that the e5 model consistently outperforms discriminative and generative models across all languages. The MPNet model follows closely, achieving results comparable to e5 in several languages. All models exhibit their best performance in English, which is expected given its prominence in the pre-training phase. Furthermore, a notable decline in effectiveness is observed when dealing with low-resource languages such as Finnish, Japanese, and Thai. Finnish, in particular, presents the most significant challenge for all models.

**Figure 2.**
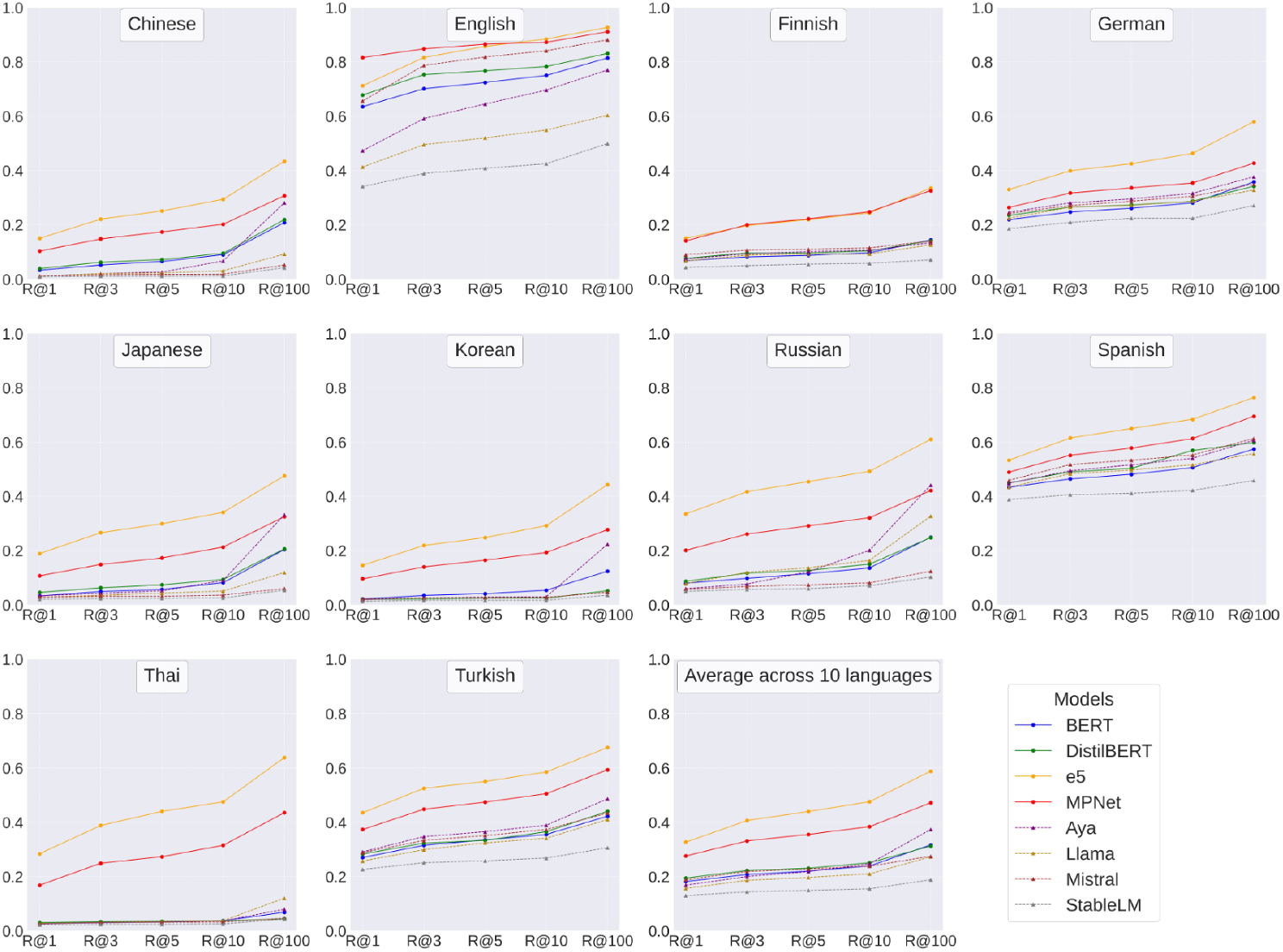
The overall performance of different models across ten languages in the XL-BEL dataset.

Language coverage may affect the performance difference between discriminative and generative LLMs. While discriminative models have been trained on over 100 languages, encompassing all target languages of our experiments, generative models do not have such extensive linguistic coverage. This could potentially explain the underperformance of generative models compared to their discriminative counterparts. However, it is worth noting that even in well-represented languages such as English, where both model types have been extensively pre-trained, generative models still fail to surpass the top-performing discriminative models. Interestingly, these results diverge from simpler contextualized biomedical concept representation in word-in-context tasks, where discriminative and generative LLMs are similar (with a small advantage for generative models) [16]. This suggests that factors beyond language coverage likely influence the performance gap between these model types.

Among generative LLMs, Mistral achieves the highest average scores. This superior performance can be attributed to its additional pre-training on biomedical corpora, which provides more extensive domain-specific knowledge. Furthermore, Mistral achieved the third-highest overall performance in English, demonstrating competitive results compared to top-performing models such as e5 and MPNet. Conversely, StableLM consistently ranks the lowest among the evaluated models, showing the weakest average performance across all 10 languages.

## 4. Discussion and Conclusion

Our comprehensive evaluation of discriminative and generative LLMs in cross-lingual biomedical concept normalization, using the XL-BEL dataset covering 10 languages, reveals several insights. Discriminative models, particularly e5 and MPNet, consistently outperformed generative models across all languages, demonstrating their efficacy in capturing the correct meaning of biomedical passages across diverse linguistic contexts.

Despite being multilingual, all models showed a significant performance decline when handling biomedical concepts in low-resource languages, highlighting a key area for improvement. Compared to other generative models, the stronger performance of Mistral, especially in English, highlights the potential of domain-specific fine-tuning for generative models in specialized tasks.

## Data Availability

All data produced in the present work are contained in the manuscript

